# Sensitivity and Uncertainty Analysis for Two-Stream Capture-Recapture Methods in Disease Surveillance

**DOI:** 10.1101/2022.09.21.22280224

**Authors:** Yuzi Zhang, Jiandong Chen, Lin Ge, John M. Williamson, Lance A. Waller, Robert H. Lyles

## Abstract

Capture-recapture methods are widely applied in estimating the number (*N*) of prevalent or cumulatively incident cases in disease surveillance. Here, we focus the bulk of our attention on the common case in which there are two data streams. We propose a sensitivity and uncertainty analysis framework grounded in multinomial distribution-based maximum likelihood, hinging on a key dependence parameter that is typically non-identifiable but is epidemiologically interpretable. Focusing on the epidemiologically meaningful parameter unlocks appealing data visualizations for sensitivity analysis and provides an intuitively accessible framework for uncertainty analysis designed to leverage the practicing epidemiologist’s understanding of the implementation of the surveillance streams as the basis for assumptions driving estimation of *N*. By illustrating the proposed sensitivity analysis using publicly available HIV surveillance data, we emphasize both the need to admit the lack of information in the observed data and the appeal of incorporating expert opinion about the key dependence parameter. The proposed uncertainty analysis is an empirical Bayes-like approach designed to more realistically acknowledge variability in the estimated *N* associated with uncertainty in an expert’s opinion about the non-identifiable parameter, together with the statistical uncertainty. We demonstrate how such an approach can also facilitate an appealing general interval estimation procedure to accompany capture-recapture methods. Simulation studies illustrate the reliable performance of the proposed approach for quantifying uncertainties in estimating *N* in various contexts. Finally, we demonstrate how the recommended paradigm has the potential to be directly extended for application to data from more than two surveillance streams.

## Introduction

The use of capture-recapture (CRC) methods has become common in epidemiological studies. Specifically, CRC methods are widely used in various disease surveillance projects for estimating total disease counts, including studies of cancer^1^, HIV infections^2-4^, COVID-19 infections^5^, and other diseases^6^.

A myriad of statistical methods have been developed for estimating the size (*N*) of a closed population. The Lincoln-Petersen (LP) estimator is the earliest example^7,8^ proposed under the independence assumption when two capture efforts are implemented. An approximately unbiased estimator was subsequently proposed by Chapman (1951)^9^ to overcome positive bias inherent in the LP estimator. In an effort to relax the independence assumption, some authors promote an estimator proposed by Chao (1987)^10-12^, which was derived under strict assumptions including many captures and small capture probabilities. For epidemiological studies involving more than two surveillance efforts, log-linear modeling has become popular because it allows certain types of dependencies between data streams^13^.

It should be noted that essentially all CRC approaches rely upon untestable assumptions to enable estimation of *N*. For example, Darroch et al. (1993)^14^ observed that the cross-product ratio measuring the dependency between two surveillance efforts cannot be estimated directly via CRC methods based on the observed data alone. Other prior studies demonstrate that the estimator of *N* can be extremely sensitive to untestable model-based assumptions, which cannot be defended using regularly-used model selection metrics^15,16^.

In epidemiological research, it is now commonplace to study the effect of untestable assumptions through sensitivity analysis in various contexts, such as assessing the effect of misclassification and unmeasured confounders^17-19^. In CRC settings, a few authors have considered sensitivity analysis to evaluate the effect of untestable assumptions on estimation of *N*^20-22^. For example, Gerritse et al. (2015)^21^ conducted a sensitivity analyses to assess the impact of a deviation from the independence assumption when estimating *N* in the two surveillance system scenario. However, given its increasing use in surveillance, CRC is a prime target for further developments toward unified and accessible sensitivity and uncertainty analyses hinging on focal parameters that are readily interpreted by epidemiologists. Doing so successfully could help to alleviate understandable doubts about the ultimate value of CRC in epidemiologic surveillance^23^.

In this work, we focus primarily on CRC experiments with two surveillance efforts. We develop a sensitivity analysis that provides a novel visualization based on a key inestimable parameter reflecting the assumed level of association between the two data streams under a population-level multinomial model^24^. In addition to exploring the sensitivity of the estimation of *N*, we further propose a simulation-based empirical Bayes-like approach to quantifying uncertainty in the estimate of *N* associated with the assumed variation in the key parameter, together with related statistical uncertainties. The proposed uncertainty analysis is a specialized example of Bayesian model averaging. While other variations on model averaging have been proposed in CRC contexts in a frequentist framework^25,26^, our proposed uncertainty analysis is similar in spirit to complex fully Bayesian analysis typically presented in literature targeting statistical audiences^27,28^. Here, we target those epidemiologists practicing surveillance by crystallizing specification of assumptions about the key parameter, offering a clear, direct and accessible approach to both sensitivity and uncertainty analyses, and demonstrating that covariates explaining heterogeneity in the population can be incorporated by stratification. We also demonstrate that special cases of the proposed uncertainty analysis can serve as a general interval estimation approach. For statistical details, we refer readers to Supplementary Materials.

## Methods

### Maximum likelihood estimators

The population-level two-stream CRC data are summarized in Table 1A. The observed number of cases with capture profile (*i, j*) is denoted as *n*_*ij*_, with subscripts of 1 indicating captured and 0 not captured by a given stream. To allow for modeling the dependency between two streams, we introduce the parameter vector 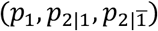 under the multinomial modeling framework, where *p*_1_ is the marginal probability of identification in stream 1, and *p*_2|1_ and 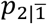 are the probability of identification by stream 2 given identified or not identified by stream 1, respectively. Among these three parameters, only *p*_2|1_ can be estimated directly from the observed data. The other two parameters are inestimable based on the data alone, since the cell count *n*_00_ is not observed. As a result of this non-identifiability, the familiar CRC modeling challenge is that at least one unverifiable assumption is required for estimating *N*.

**Table 1:** Cell Counts for Two-Stream Capture-Recapture (CRC)

| Table 1A |  |  | Table 1B<br>CRC data for monitoring HIV in Lazio, Italy,<br>during 1990<sup>a</sup> |  |  |
| --- | --- | --- | --- | --- | --- |
|  |  | Captured in Stream 2 |  |  | Captured in Stream 2 |
| Captured in Stream 1 | Yes | No | Captured in Stream 1 | Yes | No |
| Yes | $n_{11}$ | $n_{10}$ | Yes | 14 | 222 |
| No | $n_{01}$ | $n_{00} = ?$ | No | 679 | ? |

<sup>a</sup> Data from Table 2 of Abeni, Brancato, and Perucci (1994)<sup>2</sup>; the two data streams are centers I and II

Consider a situation where a researcher assumes a specific value for the inestimable parameter 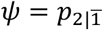. Given a valid *Ψ* (i.e., 0 < *Ψ* ≤ 1), the MLE of *N* and its variance estimator are given by^29,30^:

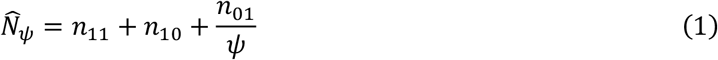

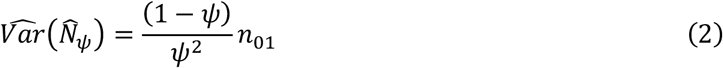

Alternatively, the unverifiable assumption can be imposed less directly via a ratio 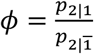. This ratio, akin to a relative risk, is a population-level measurement of dependency between the two streams. The commonly assumed Lincoln-Petersen (LP) condition (i.e., independence assumption) corresponds to *ϕ* = 1, whereas *ϕ* > 1 suggests an overall positive association between the two streams, and < 1 indicates negative association. In the CRC literature, the case of *ϕ* > 1 is referred to as “trap happiness”, while *ϕ* < 1 is known as “trap aversion”^31^. With a known population-level ratio *ϕ*, the MLE of *N* and its variance estimator become^29,30^:

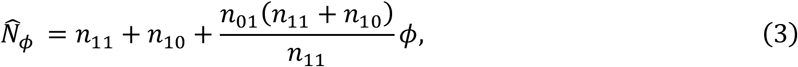

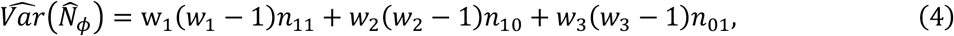

where 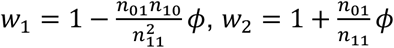, and 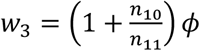. Here, Equation (4) is a simpler and more generalizable expression of a result given in Chen (2020)^29^.

The odds ratio 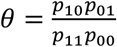 is another population-level measure of dependency that has been a focal point in previous CRC literature^14,20,32^. The parameters *ϕ* and *θ* are essentially interchangeable in that the value 1 is a benchmark for both; if *θ* indicates the two streams are positively correlated, so does *ϕ*. The closed-form MLE of *N* based on known *θ* and its variance estimator are:

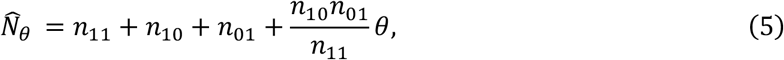

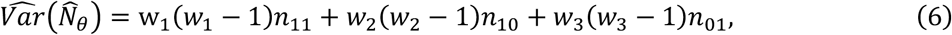

where 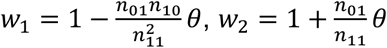, and 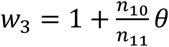. The detailed derivations for the MLEs in Equations (1), (3), and (5), along with their variances, appear in Supplementary Materials.

It is important to note that the assumptions are imposed at the population-level. For example, the assumption *ϕ* = 1 or *θ* = 1 technically allows for a mixture of individuals characterized by trap-happiness and trap-aversion, if this mixture happens to result in the state of nature that *ϕ* = 1 or *θ* = 1 at the population level^30^. Similarly, a constant value assumed for *Ψ* does not necessarily imply that this value is the same for each individual. The parameters *Ψ* and *ϕ* depend on the labeling of the data streams. Specifically, 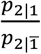 is not necessarily equal to 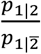 and 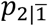 is generally different from 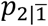. When basing sensitivity analysis on *ϕ* or *Ψ*, we suggest that researchers to start with selecting the most comfortable labeling, meaning that they are more confident in making assumptions about the key parameter under that selected labeling. In contrast, the parameter *θ* is invariant to different labeling.

The MLE of *N* given in Equation (1) is strictly unbiased, while the MLEs in Equations (3) and (5) are biased. To reduce mean bias associated with 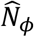, we generalize a Taylor-series expansion approach that was taken by Lyles et al. (2021)^30^ and derive two bias-corrected estimators referred to as the BC and BC2 estimators. The BC2 estimator was previously found to be nearly identical to the bias-corrected estimator of Chapman^9^ under the LP conditions^30^. Thus, one can also consider a simple direct generalization, by introducing the parameter *ϕ* into the original form of the Chapman estimator to obtain another bias-corrected estimator for a given *ϕ*; we refer to this estimator as a generalized Chapman estimator. These same bias correction procedures can be applied to 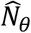 We provide derivations and algebraic forms for these bias-corrected estimators and their variances in Supplementary Materials.

### Sensitivity analysis

Unverifiable assumptions about the population-level dependency between two streams can be couched in terms of different lynchpin parameters, which can be the basis for sensitivity analysis. For example, Gerritse et al. (2015)^21^ and Zhang and Small (2020)^22^ chose the coefficient of the two-way interaction term in a log-linear model fitted to two-stream CRC data as the key parameter. Compared to the coefficient of the interaction term in a log-linear model, the parameters *Ψ, ϕ*, and *θ* are much more amenable to epidemiologic interpretation. Given that *Ψ* and *ϕ* are more easily generalizable to incorporate multiple stream case, here we focus primarily on *Ψ* or *ϕ*. We illustrate this approach with publicly available HIV surveillance data, noting that a similar sensitivity analysis could also be applied readily using *θ* as the key parameter. Motivating data were collected from four data streams in Lazio, Italy, during 1990^2^. For the purpose of illustration, we selected one pair of these data streams (centers I and II); the data are presented in Table 1B.

Figure 1 shows how the MLE of *N* varies with the assumed values of *Ψ* and *ϕ*, with point-wise Wald-type 95% confidence intervals (CI) assuming known *Ψ* or *ϕ*. lt is instructive to note how sensitive the estimated *N* is to the changes in *Ψ* and *ϕ*. The estimated *N* ranges from about 7,000 to 30,000 for *Ψ* within the range (0.02, 0.1), while *ϕ* taking values from 0.75 to 3 corresponds to the estimated *N* varying from 8,820 to 34,574. The LP estimator and the estimator proposed by Chao (1987)^10^ are labeled in the x-axis of the plots. While the LP estimator assumes *ϕ* = 1, the Chao estimator assumes a level of positive dependency estimated based on a mathematical model developed assuming conditions that would typically be unrealistic in two-stream epidemiological surveillance^30^. The LP estimator yields an estimate of 11,682 (95% CI: 5,807, 17,557); the corresponding estimated *Ψ* is 0.059. The Chao estimator yields an estimate of 29,908 (95% CI: 14,252, 45,564), with corresponding estimated *Ψ* and *ϕ* taking the values of 0.023 and 2.59, respectively. The huge difference between these two estimates of *N* stems from the vastly different projected values of *ϕ*, i.e., 1 v.s. 2.59.

**Figure 1:**
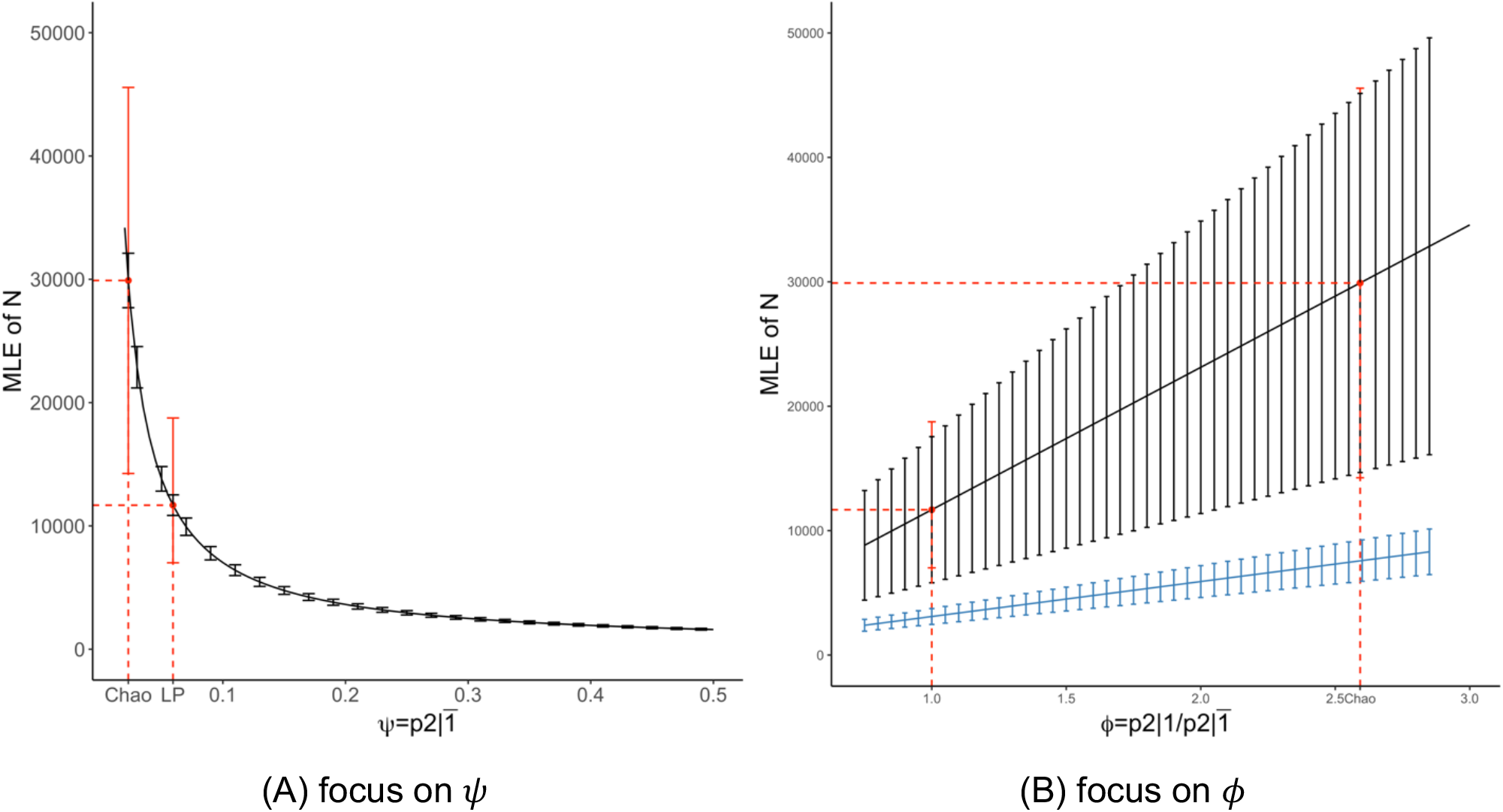
Sensitivity plots based on data from Table 2. The black error bars represent point-wise Wald-type 95% CIs assuming known *Ψ* and *ϕ*. Red solid points and error bars mark the LP estimator and the estimator of Chao (1987)^10^ along with their 95% CIs; note that in Figure 1(B), *ϕ* = 1 corresponds to the LP estimator. The blue line denotes the sensitivity plot based on the data where 50 patients in the *n*_01_ cell has moved to the *n*_11_ cell while the *n*_10_ cell remains the same.

The blue line with smaller slope in Figure 1(B) shows the sensitivity plot that would have resulted if 50 patients in the *n*_01_ cell had instead appeared in the *n*_11_ cell, with *n*_10_ remaining the same (*n*_11_ = 64, *n*_10_ = 222, *n*_01_ = 679). It is clear that based on this data, the estimation of *N* is far less sensitive to the assumption imposed on *ϕ* over the range depicted. This finding is consistent with prior observations^21^ that estimation is less sensitive when the implied coverage (measured by 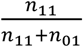) of the two streams is high.

**Table 2:**
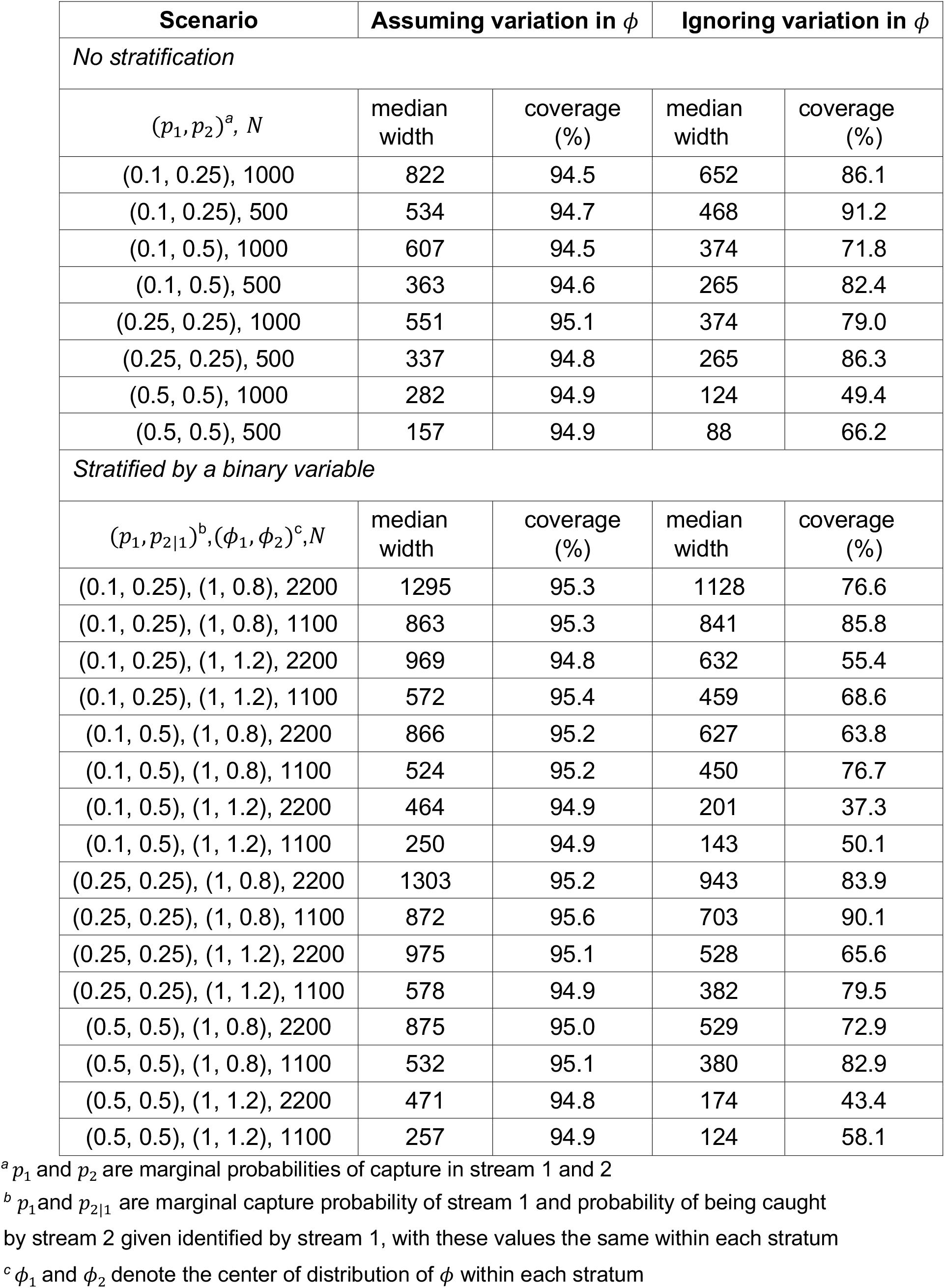
Simulation Results for Evaluating Uncertainty Quantification with and without Stratification

We emphasize that all MLEs visualized on the sensitivity plots (Figure 1) yield the exact same maximized value of the general multinomial likelihood^30^, implying that the observed data alone provide no information about what dependence assumption should be imposed for estimation. Data-driven assumptions about *Ψ* or *ϕ* obtained through metrics such as the Chao (1987) estimator or a log-linear model as applied to two streams are seldom clear to practitioners, and can also never be verified based only on the observed data^30^. This motivates us to propose an extension of the sensitivity analysis geared toward providing a fair measure of uncertainty when estimating *N* subsequent to embedding variation about *Ψ* or *ϕ* by assumption.

### Uncertainty analysis

As Figure 1 illustrates, specifying a known value for *Ψ* leads to an exceedingly precise estimator. However, this estimator can very seldom be unlocked for direct use outside of sensitivity analysis, except under a unique study design^33^. In the typical CRC scenario, we propose an uncertainty analysis anchored on the ratio parameter *ϕ*. Specifically, an assumed distribution (akin to a prior) for *ϕ* is specified beforehand. This assumed distribution would be centered at the epidemiologist’s best guess and reflects his or her level of confidence in that guess and the anticipated feasible range.

The proposed simulation-based uncertainty analysis incorporates the assumed variation in *ϕ* in a Bayes-like fashion. First, we reiterate that the data likelihood contains no information for updating *ϕ*. As such, the distribution to be postulated for *ϕ* is generally to be specified based on expert opinion. For the estimable parameter *p*_2|1_, we apply a weakly informative prior to obtain its posterior distribution as in standard Bayesian analysis.

To implement the proposed uncertainty analysis, two options are available. Option (1) is to propagate the variation in *ϕ* along with statistical uncertainties of estimable parameters into the appropriate level of uncertainty about *Ψ*, and to leverage the unbiased estimator 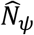 from Equation (1). Option (2) is to accommodate the variation in *ϕ* directly using a bias-corrected estimator. Detailed procedures for obtaining 95% percentile intervals for *N* under Options (1) and (2) are presented in Supplementary Materials.

As illustrated previously^30^, under the LP conditions in which *Ψ* = *p*_2|1_, adopting a Beta (1, 0) prior for *p*_2|1_ and replacing *Ψ* in Equation (1) by the resulting posterior mean yields the Chapman estimator. This connection motivates us to use the Beta (1, 0) prior for *p*_2|1_ when implementing Option (1). If implementing Option (2), we recommend use of the BC2 estimator or the generalized Chapman estimator, since the BC estimator is downward biased and suffers from instability when the probability associated with capture history (1,1) is small (Table S2).

While Option (2) can be implemented more directly, Option (1) is more easily generalizable. This is because derivations of bias-corrected estimators require extra effort and become specific to the chosen definition of the ratio parameter *ϕ* as the number of surveillance streams increases. For example, a total of three definitions of *ϕ* can be considered under three stream case (see Conclusions section and details in Supplementary Materials). In contrast, the direct analogue to 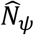 is readily available in the multiple (>2) stream case.

We conducted uncertainty analysis using Option (1), exploring different assumed distributions for *ϕ* for the HIV data (Table 1B). To promote some commonality with prior work^27^, we considered three distributions for illustration: (a) Uniform(0.75, 1.25), (b) Uniform(0.5, 1.5), and (c) Uniform(1, 2). The first two distributions reflect the same best guess, since they are both centered at 1; however, both reflect a lack of complete faith in that assumption. Compared to the first, the wider range of the second distribution indicates a lower level of confidence in the best guess of 1. The third distribution centered at 1.5 represents a case in which we assume an expert has reason to believe the two streams are positively correlated at the population level.

As shown in Figure 2, compared to assuming *ϕ* equal to a specific value, allowing variation in *ϕ* naturally results in a more conservative interval. Assuming *ϕ* = 1, the estimated *N* is the LP estimate 11,682 with 95% CI (7,165, 19,042). This CI was obtained by applying the proposed uncertainty analysis for interval estimation with the *ϕ* distribution degenerating to 1, a practice that we study empirically in a subsequent section. Taking account of uncertainty in *ϕ* results in a more conservative 95% CI. For example, the 95% CI (6,488, 20,134) was obtained based on a Uniform(0.75, 1.25) prior for *ϕ*. When the assumed distribution of *ϕ* was Uniform(1, 2), the point estimate for *N* became 17,405 (predictably higher than the LP estimate) with 95% CI (8,958, 31,054).

**Figure 2:**
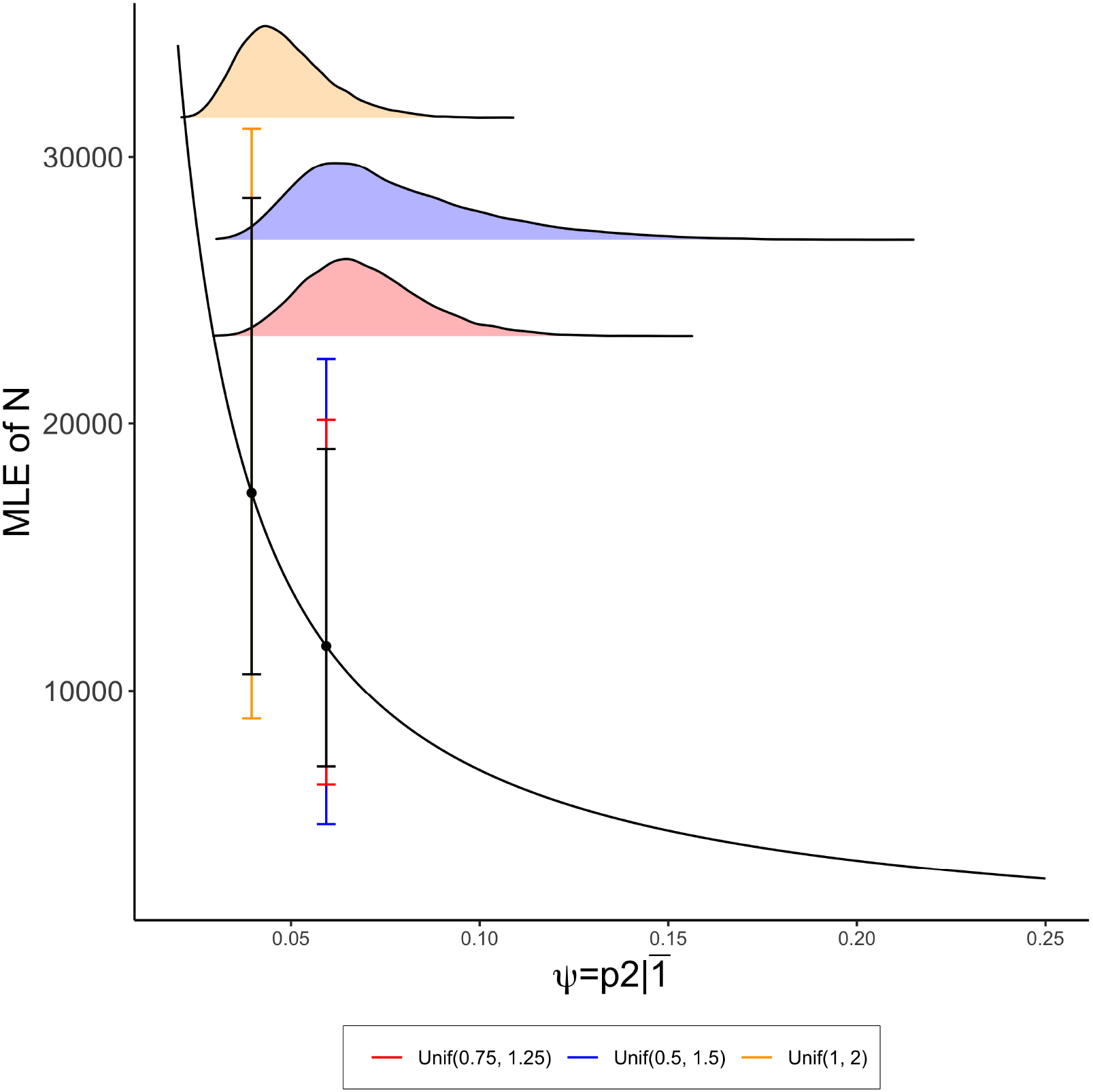
Uncertainty analysis for data from Table 2. The density plots in red, blue, and yellow reflect the posterior distributions of *Ψ* when the assumed distributions of *ϕ* are Unif(0.75, 1.25), Unif(0.5, 1.5), and Unif(1, 2), respectively. The black error bars represent the 95% CIs obtained from the proposed uncertainty analysis assuming *ϕ* = 1 (7165, 19042), and *ϕ* = 1.5 (10627, 28446). The error bars in red, blue, and yellow denote 95% CIs obtained from the uncertainty analysis with assumed distributions of *ϕ* are Unif(0.75, 1.25), Unif(0.5, 1.5), and Unif(1, 2), respectively.

We note that the LP conditions represent a natural “middle ground” for uncertainty analysis, with the LP or Chapman estimator reported with its usual measure of statistical uncertainty representing a special case in which the assumed distribution for *ϕ* degenerates to 1. This provides additional motivation to examine the proposed uncertainty analysis framework as a general approach for interval estimation of *N*. For example, when the distributional assumption for *ϕ* degenerates to a specific value, the 95% CI constructed based on the proposed uncertainty analysis can be viewed as an empirical approach to obtain interval estimation of *N* under the assumption that *ϕ* equals that specific value.

In practice, postulating the center of the assumed distribution for *ϕ* is typically a difficult task. However, expert opinion in this regard is arguably more defensible than reliance on a specific statistical model to elucidate *ϕ*, particularly in the case of two-stream disease surveillance. The task may be more readily tackled within strata formed by variables deemed to be associated with likelihood of capture at the population level. Within such strata, it may be easier for the epidemiologist to postulate where the center of the *ϕ* distribution should be, and in particular whether it should be > 1, = 1, or < 1. The choice of both the center and spread of the assumed distribution can clearly be stratum-specific. The proposed uncertainty analysis thus offers a principled way to account for covariates, acknowledging the fact that the true *ϕ* is unknown within each stratum and yielding point and interval estimates for *N* obtained by summing over strata.

### Sensitivity analysis with a known case ratio

In practice, one strategy to aid with postulating the range of key parameters is to leverage external information. For example, Wolter (1990) assumed a known ratio of cases across the sexes together with the same odds ratio (i.e., *θ*) in the male and female groups^32^ to obtain estimates of *N* and *θ*. Motivated by this idea, we apply the proposed sensitivity analysis under the assumption that the case ratio is known and that key parameters *ϕ* or *θ* are the same across strata. Specifically, suppose the targeted population is stratified by a certain binary covariate, with the proposed sensitivity analysis applied within each stratum. In this case, the crossing point of the two sensitivity plots provides a visual estimate of both *N* and the key parameter (*ϕ* or *θ*). The corresponding derived estimators are presented in Supplementary Materials. It can be shown that these estimators coincide with the MLEs of those parameters.

To visualize the information which can be directly read from the sensitivity plots anchored on *ϕ* or *θ*, we used the data explored in Wolter (1990)^32^. The dataset contains two strata (males and females) with respective sizes 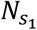 and 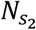, and a known sex ratio 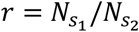 of 1.15; the data are presented in Table S1. In Figure 3(B), the crossing point corresponds to 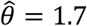, and an estimated female case count of 74, which equal the MLEs reported previously^32^. The crossing point in Figure 3(A) suggests that *ϕ* is estimated to be 1.1 and the estimated population size of females is 74. The estimated total population size *N* is 159, no matter whether we impose the equivalence assumption with respect to *ϕ* or *θ*. Although for this particular dataset the estimated *N* is the same under these two assumptions, this equivalence is not guaranteed in general. We also note that when the sensitivity plots for the two strata are parallel or overlap, the crossing point cannot be identified. In other words, the estimators of *ϕ* and *θ* are not well defined in such instances, adding a visual clarification of the potential instability in Wolter’s proposed estimator^34^.

**Figure 3:**
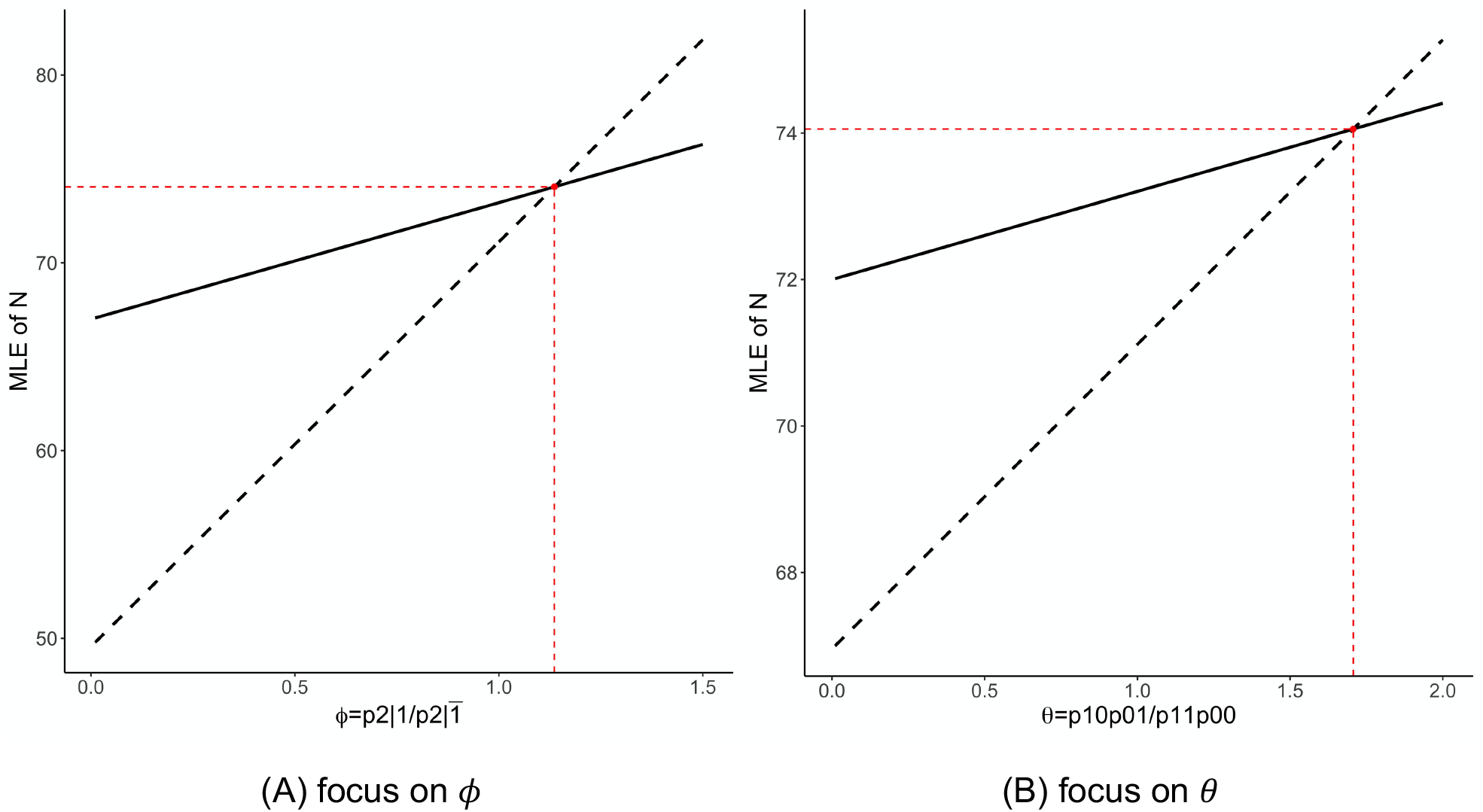
Sensitivity plot with known case ratio based on data from Table S1. The red solid points mark the crossing points of sensitivity plots; the black solid lines represent sensitivity plots for females and the black dashed lines represent scaled sensitivity plots (i.e., divided by the known sex ratio *r* = 1.15) for males.

### Simulation Studies

In this section, we summarize simulations evaluating the performance of the proposed uncertainty analysis, accounting for user-specified assumed variation in *ϕ* together with the associated statistical uncertainties. The simulation study was implemented using Option (1); results obtained under different scenarios with and without stratification are presented in Table 2. For the simulation without stratification, the data were generated assuming *ϕ* follows a Uniform(0.75, 1.25) distribution. As suggested by the coverage summary, the proposed uncertainty analysis accounting for the variation in *ϕ* fully and accurately quantified the uncertainties in estimating *N*. In contrast, failing to account for that variation by assuming a degenerate distribution for *ϕ* at 1 resulted in a severe under-coverage problem.

We also conducted a simulation study under the situation where two strata were formed. Two types of mixture were explored, the first one a mixture of independence and negative association and the second one a mixture of independence and positive association. Simulation results demonstrate that the proposed uncertainty analysis is easily adapted to incorporating stratification and providing excellent coverage when uncertainty in *ϕ* is correctly imposed (Table 2). Under-coverage is again demonstrated when variation in *ϕ* is ignored.

Finally, we conducted simulation for evaluating (1) the derived bias-corrected estimators and (2) the performance of the proposed uncertainty analysis in serving as a general interval estimation approach. The simulation results corresponding to these two purposes are displayed in Tables S2 and S3. We found that the proposed BC2 and generalized Chapman estimators are virtually unbiased, and almost identical as *N* increases. In cases where both were downward biased, the BC2 estimator was slightly less biased compared to the generalized Chapman estimator and notably less biased than the BC estimator. Interval estimation under the LP conditions based on the proposed uncertainty analysis achieved comparable performance with a previously proposed transformed logit CI^35^, both in terms of coverage and median interval width. The transformed logit CI was selected for comparison due to its reliable performance in coverage relative to numerous other methods^30,35^.

## Conclusions and Discussion

We have developed a sensitivity and uncertainty analysis framework focused upon a key inestimable parameter for CRC experiments with two surveillance efforts. Three definitions based on intuitive interpretations of that inestimable parameter are introduced, i.e., *Ψ, ϕ* and *θ*. Closed-form MLEs hinging on these key parameters are derived, where the one based on a known value of 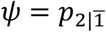 (Equation 1) was previously confirmed as strictly unbiased^30^. We have provided bias-corrected estimators as alternatives to the MLEs based on known *ϕ* and *θ* in Equations (3) and (5). The proposed sensitivity analysis can be anchored on either one of those measures of association between the two streams, to graphically study the impact of the key parameters on the estimation of *N*. Applying the proposed sensitivity analysis to motivating HIV data^2^, we have emphasized the significance of incorporating the epidemiologist’s expert opinion in terms of directionality (<1, =1, >1) and where to center the best guess for the association parameter *ϕ* or *θ*, given that the observed data alone carry no information about those parameters.

The proposed uncertainty analysis approach is further designed for quantifying uncertainties in the estimation of *N* while accounting for assumed variation in *ϕ*. While this aspect of our work is similar in spirit to fully Bayesian analyses previously developed in which estimation is carried out via dedicated algorithms^27^, our proposed approach to uncertainty analysis is much more intuitive and practical, providing general access for epidemiologic applications. Two options are available for incorporating the assumed variation of *ϕ*. Option (1) leverages the unbiased MLE assuming a given value of *Ψ*, while Option (2) directly uses a bias-corrected MLE. Choosing *Ψ* as the focus facilitates the generality of the proposed uncertainty analysis and is motivated by two points. First, the MLE assuming a known value of the parameter analogous to 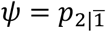 can be easily derived under the multiple stream case. Second, we find that different assumptions can cleanly be related back to the parameter analogous to *Ψ*. These two points also enable the direct extension of the proposed sensitivity analysis to incorporate multiple streams. For example, the inestimable parameter 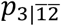 is the analogue to 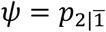 in the three stream case; we provide the MLE for *N* assuming a known 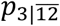 (the analogue to Equation 1) in Supplementary Materials. As part of ongoing work, we aspire to fully develop the extension to multiple streams, and to use the methods provided here to motivate a transparent general modeling framework for analyzing CRC data in epidemiological surveillance studies. In the current article, we have also illustrated that the proposed uncertainty analysis can serve as a general interval estimation approach with reliable performance for CRC experiments under various scenarios.

As noted, the key association parameter cannot be estimated using observed data alone, and we acknowledge that this non-identifiability precludes any complete solution to the CRC problem (except under very specific study designs^33^). We emphasize that most CRC methods used in practice, especially in the case of two streams, essentially ignore this identifiability problem. This highlights a pressing need for sensitivity and uncertainty analyses to leverage expert opinion to whatever extent possible to drive the dependence assumption upon which estimation hinges. Focusing on key dependence parameters (*Ψ,ϕ*, and *θ*) that are amenable to epidemiologic interpretation provides accessibility and practical value that are arguably lacking in prior attempts to promote sensitivity analysis for CRC.

## Data Availability

All data produced in the present work are contained in the manuscript

## Acknowledgments

We thank Drs. Howard Chang and Sarita Shah for motivation and helpful discussions. Partial support was provided by the National Institute of Health-funded Emory Center for AIDS Research (P30AI050409; Del Rio PI), the National Center for Advancing Translational Sciences of the National Institutes of Health (UL1TR002378; Taylor PI) and by the National Institutes of Health/National Cancer Institute-funded Cancer Recurrence Information and Surveillance Program (CRISP) study (1 R01 CA208367-01; Ward/Lash MPIs). The content is the sole responsibility of the authors and does not necessarily represent the official views of the National Institutes of Health.

## Notes

### Competing Interest Statement

The authors have declared no competing interest.

### Author Declarations

Data used in this paper are publicly available data

