## Supplementary Materials for "Sensitivity and Uncertainty Analysis for Two-Stream Capture-Recapture Methods in Disease Surveillance"

### A. Multinomial Model

$$(N_{11}, N_{10}, N_{01} | N_c = n_c) \sim \text{Multinomial}(n_c, p_{11}^*, p_{10}^*, p_{01}^*),$$

where $n_c = n_{11} + n_{10} + n_{01}$ is the number of cases caught at least once; $p_{ij}^* = p_{ij}/p_c$ in which $p_c = p_{11} + p_{10} + p_{01}$ and $p_{ij}$ denotes the probability of having capture history $(i,j)$, $i,j \in \{0,1\}$.

$$L(p_c) = \frac{n_c}{n_{11}!\, n_{10}!\, n_{01}!} \left(\frac{p_{11}}{p_c}\right)^{n_{11}} \times \left(\frac{p_{10}}{p_c}\right)^{n_{10}} \times \left(\frac{p_{01}}{p_c}\right)^{n_{01}}$$

$$\widehat{N} = \frac{n_c}{\hat{p}_c}$$

### B. Derivation of Bias-Corrected Estimators

Given $\phi$, the MLE of $N$ is

$$\widehat{N}_\phi = n_{11} + n_{10} + \frac{n_{01}(n_{11} + n_{10})}{n_{11}} \phi$$

Define $n_c = n_{11} + n_{10} + n_{01}, p_c = p_{11} + p_{10} + p_{01}$ and $p_{ij}^* = \frac{p_{ij}}{p_c}$, where $i,j \in \{0,1\}$. Dividing both sides by $\widehat{N}_\phi$, we have:

$$1 = \hat{p}_{11} + \hat{p}_{10} + \frac{\hat{p}_{01}(\hat{p}_{11} + \hat{p}_{10})}{\hat{p}_{11}} \phi,$$

$$\frac{1}{\hat{p}_c} = \hat{p}_{11}^* + \hat{p}_{10}^* + \frac{\hat{p}_{01}^*(\hat{p}_{11}^* + \hat{p}_{10}^*)}{\hat{p}_{11}^*} \phi.$$

Then, the $\widehat{N}_\phi$ can be written as:

$$\widehat{N}_\phi = n_{11} + n_{10} + \frac{n_{01}(n_{11} + n_{10})}{n_{11}} \phi = n_c \left\{ \hat{p}_{11}^* + \hat{p}_{10}^* + \frac{\hat{p}_{01}^*(\hat{p}_{11}^* + \hat{p}_{10}^*)}{\hat{p}_{11}^*} \phi \right\} = \frac{n_c}{\hat{p}_c}.$$

Let $\boldsymbol{p} = (p_{11}^*, p_{10}^*, p_{01}^*)$ and define a function $f(\widehat{\boldsymbol{p}}^*) = \hat{p}_{11}^* + \hat{p}_{10}^* + \frac{\hat{p}_{01}^*(\hat{p}_{11}^* + \hat{p}_{10}^*)}{\hat{p}_{11}^*} \phi$. Follow the Taylor-series expansion[1]:

$$E[f(\widehat{\boldsymbol{p}}^*)] = f(\widehat{\boldsymbol{p}}^*) + g(\widehat{\boldsymbol{p}}^*) + O(n_c^{-2}),$$

where $g(\boldsymbol{p}^*) = E[\frac{1}{2}(\widehat{\boldsymbol{p}}^* - \boldsymbol{p}^*)^T \boldsymbol{D}_2(\boldsymbol{p}^*)(\widehat{\boldsymbol{p}}^* - \boldsymbol{p}^*)]$, and $\boldsymbol{D}_2(\boldsymbol{p}^*)$ is the Hessian of $f$ evaluated at $\boldsymbol{p}^*$, which is given by

$$\begin{bmatrix} \frac{2p_{10}^* p_{01}^*}{(p_{11}^*)^3}\phi & -\frac{p_{01}^*\phi}{(p_{11}^*)^2} & -\frac{p_{10}^*\phi}{(p_{11}^*)^2} \\ -\frac{p_{01}^*\phi}{(p_{11}^*)^2} & 0 & \frac{\phi}{p_{11}^*} \\ -\frac{p_{10}^*\phi}{(p_{11}^*)^2} & \frac{\phi}{p_{11}^*} & 0 \end{bmatrix}$$

Then we have $g(\boldsymbol{p}^*)$ is:

$g(\boldsymbol{p}^*) = \frac{1}{2} \times \frac{2p_{10}^* p_{01}^*}{(p_{11}^*)^3} Var(\hat{p}_{11}^*) + \frac{-p_{01}^* \phi}{(p_{11}^*)^2} Cov(\hat{p}_{11}^* + \hat{p}_{10}^*) + \frac{-p_{10}^* \phi}{(p_{11}^*)^2} Cov(\hat{p}_{11}^* + \hat{p}_{01}^*) + \frac{\phi}{p_{11}^*} Cov(\hat{p}_{10}^*, \hat{p}_{01}^*)$.

Under the conditional multinomial distribution model, we have:

$Var(\hat{p}_{11}^*) = \frac{p_{11}^*(1-p_{11}^*)}{n_c}, Cov(\hat{p}_{11}^*, \hat{p}_{10}^*) = \frac{-p_{11}^* p_{10}^*}{n_c}, Cov(\hat{p}_{11}^*, \hat{p}_{01}^*) = \frac{-p_{11}^* p_{01}^*}{n_c}, Cov(\hat{p}_{10}^*, \hat{p}_{01}^*) = -\frac{p_{10}^* p_{01}^*}{n_c}$.

Some algebra leads to:

$$\hat{g}(\boldsymbol{p}^*) = \frac{n_{10} n_{01} \phi}{n_{11}^2 n_c}.$$

Since $\widehat{N}_\phi = n_c f(\boldsymbol{p}^*)$, the estimated bias in $\widehat{N}_\phi$ is:

$$n_c \hat{g}(\boldsymbol{p}^*) = \frac{n_{10} n_{01}}{n_{11}^2} \phi.$$

Finally, the bias-corrected (BC) estimator of $N$ and its variation estimator are given by

$$\widehat{N}_\phi^{BC} = \widehat{N}_\phi - \frac{n_{10} n_{01}}{n_{11}^2} \phi. \qquad (S1)$$

$$\widehat{Var}\left(\widehat{N}_\phi^{BC}\right) = \mathrm{w}_1(w_1 - 1 - C) n_{11} + w_2(w_2 - 1 - C) n_{10} + w_3(w_3 - 1 - C) n_{01}, \qquad (S2)$$

where $C = \frac{n_{10} \hat{p}_{01}}{n_{11}^2} \phi$, $w_1 = 1 - \frac{n_{10} n_{01}}{n_{11}^2} \phi + \frac{2 n_{01} n_{10}}{n_{11}^3} \phi$, $w_2 = 1 + \frac{n_{01}}{n_{11}} \phi - \frac{n_{01}}{n_{11}^2} \phi$, and $w_3 = \phi + \frac{n_{10}}{n_{11}} \phi - \frac{n_{10}}{n_{11}^2} \phi$.

It is clear that the correction term is not well-defined when $n_{11} = 0$. A small adjustment to the denominator term in the correction factor results in:

$$\widehat{N}_\phi^{BC2} = \widehat{N}_\phi - \frac{n_{10} n_{01}}{(n_{11} + 0.5)^2} \phi. \qquad (S3)$$

The variance estimator of the estimator $\widehat{N}_\phi^{BC2}$ derived using the multivariate delta method is:

$$\widehat{Var}\left(\widehat{N}_\phi^{BC2}\right) = \mathrm{w}_1(w_1 - 1 - C) n_{11} + w_2(w_2 - 1 - C) n_{10} + w_3(w_3 - 1 - C) n_{01}, \qquad (S4)$$

where $C = \frac{2 n_{11} n_{01} \hat{p}_{01}}{(n_{11}+0.5)^3} - \frac{n_{10} \hat{p}_{01}}{(n_{11}+0.5)^2} \phi$, $w_1 = 1 - \frac{n_{10} n_{01}}{n_{11}^2} \phi + \frac{2 n_{10} n_{01}}{(n_{11}+0.5)^3} \phi$, $w_2 = 1 + \frac{n_{01}}{n_{11}} \phi - \frac{n_{01}}{(n_{11}+0.5)^2} \phi$, and $w_3 = \phi + \frac{n_{10}}{n_{11}} \phi - \frac{n_{10}}{(n_{11}+0.5)^2} \phi$.

The estimator $\widehat{N}_\phi^{BC}$ is a direct generalization of the corrected MLE of Darroch (1958)[2] in the two-stream case under the LP conditions, while $\widehat{N}_\phi^{BC2}$ (BC2 estimator) generalizes an estimator shown by Lyles et al. (2021)[3] to be a direct competitor to the bias-corrected estimator of Chapman (1951)[4]. One can also consider a simple direct generalization, by introducing the parameter $\phi$ into the original form of the Chapman estimator, as follows:

$$\widehat{N}_{Chap}^* = \frac{(n_{11} + n_{10} + 1)(n_{11} + n_{01}\phi + 1)}{(n_{11} + 1)} - 1 \qquad (S5)$$

Note that Equation (S5) reduces to the original Chapman estimator when $\phi = 1$, and we refer it as the generalized Chapman estimator. The corresponding variance estimator is:

$$\widehat{Var}\left(\widehat{N}^{*}_{Chap}\right) = \mathrm{w}_1(w_1 - 1 - C)n_{11} + w_2(w_2 - 1 - C)n_{10} + w_3(w_3 - 1 - C)n_{01}, \quad (S6)$$

where $C = \frac{n_{10}\hat{p}_{01}}{(n_{11}+1)^2}\phi$, $w_1 = 1 - \frac{n_{10}n_{01}}{(n_{11}+1)^2}\phi$, $w_2 = 1 + \frac{n_{01}}{n_{11}+1}\phi$, and $w_3 = \phi + \frac{n_{10}}{n_{11}+1}\phi$.

Analogous to the recommendations of Lyles et al. (2021)[3], we suggest setting $\widehat{N}^{BC2}_{\phi}$ in (S3) equal to $\widehat{N}^{*}_{Chap}$ in (S5), in the event that $n_{11} = 0$ when using the BC2 approach.

The Taylor-series expansion approach can also be applied to bias-correct the MLE with known odds ratio $\theta$. We first write the MLE of $N$ in Equation (5) as:

$$\widehat{N}_{\theta} = \frac{n_c}{\hat{p}^{*}_{11} + \hat{p}^{*}_{10} + \hat{p}^{*}_{01} + \frac{\hat{p}^{*}_{10}\hat{p}^{*}_{01}}{\hat{p}^{*}_{11}}\theta}$$

Similarly, we define $f(\widehat{\boldsymbol{p}}^{*}) = \hat{p}^{*}_{11} + \hat{p}^{*}_{10} + \hat{p}^{*}_{01} + \frac{\hat{p}^{*}_{10}\hat{p}^{*}_{01}}{\hat{p}^{*}_{11}}\theta$, and $\boldsymbol{D}_2(\boldsymbol{p}^{*})$ evalued at $\boldsymbol{p}^{*}$ is

$$\begin{bmatrix} \frac{2p^{*}_{10}p^{*}_{01}}{(p^{*}_{11})^3}\theta & -\frac{p^{*}_{01}\theta}{(p^{*}_{11})^2} & -\frac{p^{*}_{10}\theta}{(p^{*}_{11})^2} \\ -\frac{p^{*}_{01}\theta}{(p^{*}_{11})^2} & 0 & \frac{\theta}{p^{*}_{11}} \\ -\frac{p^{*}_{10}\theta}{(p^{*}_{11})^2} & \frac{\theta}{p^{*}_{11}} & 0 \end{bmatrix}$$

Some algebra leads to:

$$\hat{g}(\boldsymbol{p}^{*}) = \frac{n_{10}n_{01}\theta}{n^2_{11}n_c}$$

The bias-corrected estimator with given $\theta$ is given by

$$\widehat{N}^{BC}_{\theta} = \widehat{N}_{\theta} - \frac{n_{10}n_{01}\theta}{n^2_{11}} \quad (S7)$$

As with the BC2 estimator with given $\phi$ in Equation (S3), the same adjustment can also be applied to stabilize the estimator in Equation (S7). The corresponding BC2 estimator with given $\theta$ is

$$\widehat{N}^{BC2}_{\theta} = \widehat{N}_{\theta} - \frac{n_{10}n_{01}\theta}{(n_{11} + 0.5)^2} \quad (S8)$$

### C. Variance Estimators

For notational convenience, we let $\boldsymbol{n}$ denote the vector of observed counts $(n_{11}, n_{10}, n_{01})$ and write the MLE of $N$ with a given value of a key parameter (e.g., $\psi$, $\phi$, or $\theta$) as $\widehat{N} = f(\boldsymbol{n})$. The first derivative of $\widehat{N}$ with respect to $\boldsymbol{n}$ is denoted as $(w_1, w_2, w_3)$, with a corresponding variance estimator derived using the multivariate delta method given by

$$\begin{aligned} Var\left(\widehat{N}\right) = N\{ & w_1p_{11}[w_1(1 - p_{11}) - w_2p_{10} - w_3p_{01}] \\ & + w_2p_{10}[w_2(1 - p_{10}) - w_1p_{11} - w_3p_{01}] \\ & + w_3p_{01}[w_3(1 - p_{01}) - w_1p_{11} - w_2p_{10}]\} \end{aligned} \quad (S9)$$

1. Variance estimator of $\widehat{N}_{\psi}$:

Some algebra based on the fact that $p_{11} + p_{10} + \frac{p_{01}}{\psi}$ = 1 leads to the variance estimator in Equation (2).

2. Variance estimator of $\widehat{N}_{\phi}$:

Some algebra based on the fact that $p_{11} + p_{10} + p_{01}\phi + \frac{p_{10}p_{01}}{p_{11}}\phi$ = 1 leads to the variance estimator in Equation (4).

3. Variance estimator of $\widehat{N}_{\phi}^{BC}$:

Some algebra based on the fact that $p_{11} + p_{10} + \left(p_{01} + \frac{p_{10}p_{01}}{p_{11}}\right)\phi - \frac{p_{10}p_{01}}{p_{11}n_{11}}\phi$ = 1 leads to the variance estimator in Equation (S2).

4. Variance estimator of $\widehat{N}_{\phi}^{BC2}$:

Some algebra based on the fact that $p_{11} + p_{10} + \left(p_{01} + \frac{p_{10}p_{01}}{p_{11}}\right)\phi - \frac{p_{10}p_{01}n_{11}}{p_{11}(n_{11}+0.5)^2}\phi$ = 1 leads to the variance estimator in Equation (S4).

5. Variance estimator of $\widehat{N}_{chap}^{*}$:

Some algebra based on the fact that $p_{11} + p_{10} + p_{01}\phi + \frac{p_{10}p_{01}n_{11}}{p_{11}(n_{11}+1)}\phi$ = 1 leads to the variance estimator in Equation (S6).

### D. Procedure for Obtaining 95% Percentile Interval for $N$

Under Option (1), the recommended procedure for obtaining a 95% percentile interval for $N$ (from which we seek favorable frequentist properties) is:

(i). Specify the prior distribution of $p_{2|1}$ and the assumed distribution of $\phi$.

(ii). Obtain $D$ posterior samples of $\psi$ by combining $D$ posterior samples of $p_{2|1}$ and $D$ independent random draws of $\phi$ generated from the assumed distribution using the definition $\psi = \frac{p_{2|1}}{\phi}$ (the posterior samples for $\psi$ empirically reflect both the assumed variation in $\phi$ and statistical uncertainty in estimating $p_{2|1}$).

(iii). For each generated $\psi$, take $M$ random draws from a $Normal(\widehat{N}_{\psi}, \widehat{Var}(\widehat{N}_{\psi}))$ distribution (see Equations (1) and (2)).

(iv). A total of $D \times M$ posterior samples of $N$ are obtained by combining together all random draws across each generated value of $\psi$.

(v). Take the 2.5$^{th}$ and 97.5$^{th}$ percentiles of the resulting $D \times M$ posterior samples of $N$ to construct the 95% percentile interval.

With a Beta (1,0) prior for $p_{2|1}$, the conjugate posterior distribution of $p_{2|1}$ is a Beta distribution with mean equal to $\frac{n_{11}+1}{n_{11}+n_{10}+1}$. Lyles et al. (2021)[3] show that inserting this mean into Equation (1) in place of $\psi$ yields the Chapman estimator. With this in mind, we recommend use of the Beta (1,0) prior for $p_{2|1}$ when implementing the proposed uncertainty analysis under Option (1).

The 95% credible interval for $N$ obtained under Option (2) is produced as follows:

(i). Specify the prior distribution of $\phi$ and generate $D$ realizations from that distribution. Importantly, accept realizations only if they yield a value greater than $\frac{n_{11}}{n_{11}+n_{10}}$; this is because $\phi = \frac{p_{2|1}}{\psi}$ and $\psi$ is constrained to be $\leq 1$.

(ii). For each given value of $\phi$, draw $M$ random draws from a normal distribution with mean $\widehat{N}_{\phi}^{BC2}$ and variance equal to the estimated variance of $\widehat{N}_{\phi}^{BC2}$ (note that $\widehat{N}_{\phi}^{BC2}$ could be replaced by any of the bias-corrected estimators in Equations (S1), (S3), and (S5)).

(iii). Take the 2.5th and 97.5th percentiles of the resulting $D \times M$ posterior samples of $N$ to construct the 95% credible interval.

### E. Crossing Points of Sensitivity Plots Obtained from Two Strata

Let $n_{11}^{s_1}, n_{10}^{s_1}$, and $n_{01}^{s_1}$ denote the observed cell counts in the first stratum, with the superscript $s_2$ used analogously for the data in the second stratum. Letting $N_{s_1}$ and $N_{s_2}$ represent the true number of cases for each stratum, the case ratio $r = N_{s_1}/N_{s_2}$ is assumed known (e.g., obtained from previous data on prevalence). Considering the proposed sensitivity analysis focused on $\phi$, it can be shown that the x-axis and y-axis coordinates of the crossing point between the plots from stratum 2 and the scaled plots from stratum 1 (i.e., depicting $N_{s_1}$ divided by $r$) are:

$$\hat{\phi} = \frac{\frac{\left(n_{11}^{s_1} + n_{10}^{s_1}\right)}{r} - \left(n_{11}^{s_2} + n_{10}^{s_2}\right)}{\frac{n_{01}^{s_2}\left(n_{11}^{s_2} + n_{10}^{s_2}\right)}{n_{11}^{s_2}} - \frac{n_{01}^{s_1}\left(n_{11}^{s_1} + n_{10}^{s_1}\right)}{r n_{11}^{s_1}}} \qquad (S10)$$

$$\widehat{N}_{s_2,\hat{\phi}} = n_{11}^{s_2} + n_{10}^{s_2} + \frac{n_{01}^{s_2}\left(n_{11}^{s_2} + n_{10}^{s_2}\right)}{n_{11}^{s_2}}\hat{\phi} \qquad (S11)$$

Similarly, the crossing point of the two sensitivity analysis plots anchored on $\theta$ are given by

$$\hat{\theta} = \frac{r\left(n_{11}^{s_2} + n_{10}^{s_2} + n_{01}^{s_2}\right) - \left(n_{11}^{s_1} + n_{10}^{s_1} + n_{01}^{s_1}\right)}{\frac{n_{10}^{s_1} n_{01}^{s_1}}{n_{11}^{s_1}} - \frac{r n_{10}^{s_2} n_{01}^{s_2}}{n_{11}^{s_2}}} \qquad (S12)$$

$$\widehat{N}_{s_2,\hat{\theta}} = n_{11}^{s_2} + n_{10}^{s_2} + n_{01}^{s_2} + \frac{n_{10}^{s_2} n_{01}^{s_2}}{n_{11}^{s_2}}\hat{\theta} \qquad (S13)$$

### F. MLEs with a Known Case Ratio

Let $n_c^{s_1}$ and $n_c^{s_2}$ denote the number of distinct cases identified by the two data streams for each stratum. The $p_{11}^{s_1}, p_{10}^{s_1}$, and $p_{01}^{s_1}$ denote corresponding capture probabilities for the first stratum and the probability of being caught at least once for the first stratum is denoted as $p_c^{s_1} = p_{11}^{s_1} + p_{10}^{s_1} + p_{01}^{s_1}$. Similarly, the superscript $s_2$ is used for the second stratum. The conditional multinomial likelihood is given by

$$L = \frac{n_c^{s_1}!}{n_{11}^{s_1}!\, n_{10}^{s_1}!\, n_{01}^{s_1}!}\left(p_{11}^{s_1}\right)^{n_{11}^{s_1}}\left(p_{10}^{s_1}\right)^{n_{10}^{s_1}}\left(p_{01}^{s_1}\right)^{n_{01}^{s_1}} \frac{n_c^{s_2}!}{n_{11}^{s_2}!\, n_{10}^{s_2}!\, n_{01}^{s_2}!}\left(p_{11}^{s_2}\right)^{n_{11}^{s_2}}\left(p_{10}^{s_2}\right)^{n_{10}^{s_2}}\left(p_{01}^{s_2}\right)^{n_{01}^{s_2}}$$

The MLEs of $p_c^{s_1}$ and $p_c^{s_2}$ under the assumption that $\phi$ is the same across strata have the following forms:

$$\hat{p}_c^{s_1} = \frac{n_c^{s_1}}{n_c^{s_1} + \frac{n_{10}^{s_1} n_{01}^{s_1}}{n_{11}^{s_1}}\phi} \qquad (S14)$$

$$\hat{p}_c^{s_2} = \frac{n_c^{s_2}}{n_c^{s_2} + \frac{n_{10}^{s_2} n_{01}^{s_2}}{n_{11}^{s_2}}\phi} \quad (S15)$$

Since we have $\frac{n_c^{s_1}}{p_c^{s_1}} = \frac{rn_c^{s_2}}{p_c^{s_2}}$, the MLE of $\phi$ is

$$\hat{\phi} = \frac{\frac{n_{11}^{s_1} + n_{10}^{s_1}}{r} - \left(n_{11}^{s_2} + n_{10}^{s_2}\right)}{\frac{n_{01}^{s_2}\left(n_{11}^{s_2} + n_{10}^{s_2}\right)}{n_{11}^{2}} - \frac{n_{01}^{s_1}\left(n_{11}^{s_1} + n_{10}^{s_1}\right)}{rn_{11}^{s_1}}} \quad (S16)$$

Supplying the MLE of $\phi$ to Equation (S15) and using the fact that $N_{s_2} = \frac{n_c^{s_2}}{p_c^{s_2}}$, the MLE of $N_{s_2}$ is

$$\hat{N}_{s_2,\hat{\phi}} = n_{11}^{s_2} + n_{10}^{s_2} + \frac{n_{01}^{s_2}\left(n_{11}^{s_2} + n_{10}^{s_2}\right)}{n_{11}^{s_2}}\hat{\phi} \quad (S17)$$

The MLEs of $\phi$ and $N_{s_2}$coincide with estimators in Equations (S10) and (S11). We also verified that estimators in Equations (S12) and (S13) are the same as the MLEs provided in Wolter (1990)[5] (see Equations 4 and 5 in Wolter 1990) derived using multinomial model.

#### G. MLEs under Three-Stream CRC

When three overlapping surveillance streams are implemented, 7 observed cell counts are obtained, denoted as $n_{111}, n_{110}, n_{101}, n_{100}, n_{011}, n_{010}, n_{001}$. The MLE and its variance estimator with given $p_{3|\overline{12}}$ derived based on population-level multinomial model are given by

$$\hat{N} = n_{111} + n_{110} + n_{101} + n_{100} + n_{011} + n_{010} + \frac{n_{001}}{p_{3|\overline{12}}} \quad (S18)$$

$$\widehat{Var}(\hat{N}) = \frac{\left(1 - p_{3|\bar{1}\bar{2}}\right)}{(p_{3|\bar{1}\bar{2}})^2} n_{001} \quad (S19)$$

Note that Equation (S18) is a direct generalization of Equation (1). Three inestimable ratio parameters (akin to relative risks) are available as focal points for sensitivity analysis in this case, i.e., $\phi_1 = \frac{p_{3|12}}{p_{3|\overline{12}}}$, $\phi_2 = \frac{p_{3|1\bar{2}}}{p_{3|\overline{12}}}$, and $\phi_3 = \frac{p_{3|\bar{1}2}}{p_{3|\overline{12}}}$. Any of the three ratio parameters would have an intuitive interpretation, for example, the ratio assumption $\frac{p_{3|1\bar{2}}}{p_{3|\overline{12}}} = 1$ reflects an assumption that identification in stream 3 is not impacted by identification in stream 1, given non-identification in stream 2.

Table S1: Cell Counts for Two-Stream Capture-Recapture Analyzed in Wolter (1990)[5]

| Males | | | Females | | |
|---|---|---|---|---|---|
| | Captured in Stream 2 | | | Captured in Stream 2 | |
| Captured in Stream 1 | Yes | No | Captured in Stream 1 | Yes | No |
| Yes | 46 | 11 | Yes | 54 | 13 |
| No | 20 | ? | No | 5 | ? |

Table S2: Simulation Results for Evaluating Bias-Corrected Estimators of $N$ with a Known $\phi$

| **Scenario** | **Mean (SD)**[a] | | | |
|---|---|---|---|---|
| $(p_1, \psi, \phi)^b$, $N$ | $\widehat{N}_{\phi}$ | $\widehat{N}_{\phi}^{BC}$ | $\widehat{N}_{\phi}^{BC2}$ | $\widehat{N}_{chap}^{*}$ |
| (0.1, 0.2, 0.75), 50 | - | 27 (18) | 33 (16) | 32 (16) |
| (0.1, 0.3, 0.75), 50 | - | 31 (22) | 40 (19) | 39 (19) |
| (0.1, 0.2, 1), 50 | - | 30 (21) | 38 (19) | 37 (19) |
| (0.1, 0.3, 1), 50 | 64 (44) | 34 (23) | 44 (22) | 43 (22) |
| (0.1, 0.2, 1.5), 50 | 64 (46) | 34 (24) | 44 (24) | 43 (23) |
| (0.1, 0.3, 1.5), 50 | 64 (45) | 39 (22) | 48 (23) | 47 (23) |
| (0.1, 0.2, 0.75), 100 | 134 (88) | 61 (49) | 85 (43) | 83 (43) |
| (0.1, 0.3, 0.75), 100 | 138 (99) | 69 (49) | 95 (49) | 92 (47) |
| (0.1, 0.2, 1), 100 | 139 (100) | 66 (51) | 93 (49) | 90 (48) |
| (0.1, 0.3, 1), 100 | 134 (99) | 77 (45) | 99 (50) | 97 (48) |
| (0.1, 0.2, 1.5), 100 | 134 (101) | 78 (47) | 99 (52) | 97 (50) |
| (0.1, 0.3, 1.5), 100 | 120 (76) | 90 (34) | 101 (42) | 100 (40) |
| (0.1, 0.2, 0.75), 200 | 284 (219) | 145 (100) | 198 (107) | 193 (102) |
| (0.1, 0.3, 0.75), 200 | 256 (183) | 171 (76) | 201 (96) | 199 (91) |
| (0.1, 0.2, 1), 200 | 265 (204) | 163 (86) | 201 (104) | 198 (99) |
| (0.1, 0.3, 1), 200 | 236 (143) | 187 (65) | 201 (82) | 200 (79) |
| (0.1, 0.2, 1.5), 200 | 235 (143) | 187 (66) | 201 (84) | 200 (81) |
| (0.1, 0.3, 1.5), 200 | 215 (72) | 197 (48) | 200 (54) | 200 (54) |
| (0.1, 0.2, 0.75), 350 | 437 (302) | 311 (126) | 352 (163) | 349 (156) |
| (0.1, 0.3, 0.75), 350 | 396 (188) | 338 (95) | 352 (122) | 351 (118) |
| (0.1, 0.2, 1), 350 | 406 (226) | 332 (106) | 351 (137) | 350 (132) |
| (0.1, 0.3, 1), 350 | 378 (121) | 347 (87) | 351 (95) | 351 (94) |
| (0.1, 0.2, 1.5), 350 | 377 (123) | 346 (90) | 350 (97) | 350 (97) |
| (0.1, 0.3, 1.5), 350 | 363 (76) | 349 (67) | 350 (68) | 350 (68) |

[a] Mean and SD denote averaged bias and standard deviation of point estimates across 10,000 simulations for each estimator, respectively; $\widehat{N}_{\phi}$ is not reported if more than 30% of simulated datasets had $n_{11}$= 0.

[b] $p_1$, $\psi = p_{2|\bar{1}}$, and $\phi = p_{2|1}/p_{2|\bar{1}}$ are probabilities used for generating data based on multinomial distribution.

Table S3: Simulation Results for Evaluating Interval Estimation Under Lincoln-Petersen Conditions

| **Scenario** | **Uncertainty analysis** | | | **Transformed logit** | | |
|---|---|---|---|---|---|---|
| $(p_1, p_2)^a$, $N$ | median width | coverage (%) | (% missed high, % missed low) | median width | coverage (%) | (% missed high, % missed low) |
| (0.05, 0.05), 10000 | 7592 | 95.0 | (1.91, 3.09) | 7499 | 95.3 | (1.36, 3.38) |
| (0.05, 0.05), 5000 | 5506 | 95.1 | (1.65, 3.25) | 5382 | 95.6 | (0.90, 3.49) |
| (0.05, 0.05), 2500 | 4042 | 95.2 | (1.12, 3.65) | 3869 | 95.9 | (0.11, 4.03) |
| (0.05, 0.1), 10000 | 5161 | 95.1 | (2.00, 2.86) | 5146 | 95.2 | (1.81, 3.00) |
| (0.05, 0.1), 5000 | 3703 | 94.9 | (1.79, 3.28) | 3678 | 95.2 | (1.48, 3.31) |
| (0.05, 0.1), 2500 | 2638 | 95.2 | (1.52, 3.25) | 2606 | 95.7 | (0.90, 3.38) |
| (0.05, 0.2), 10000 | 3430 | 94.7 | (2.25, 3.03) | 3434 | 94.8 | (2.23, 3.00) |
| (0.05, 0.2), 5000 | 2439 | 94.8 | (2.27, 2.94) | 2437 | 95.1 | (2.11, 2.80) |
| (0.05, 0.2), 2500 | 1744 | 95.0 | (1.76, 3.26) | 1742 | 95.3 | (1.61, 3.13) |
| (0.1, 0.1), 10000 | 3534 | 95.2 | (1.99, 2.83) | 3535 | 95.1 | (1.93, 2.93) |
| (0.1, 0.1), 5000 | 2519 | 94.8 | (2.22, 3.02) | 2514 | 95.1 | (1.99, 2.91) |
| (0.1, 0.1), 2500 | 1797 | 95.1 | (1.83, 3.08) | 1788 | 95.4 | (1.58, 2.98) |
| (0.1, 0.2), 10000 | 2356 | 95.0 | (2.15, 2.88) | 2358 | 95.1 | (2.03, 2.89) |
| (0.1, 0.2), 5000 | 1669 | 94.8 | (2.31, 2.85) | 1669 | 95.1 | (2.24, 2.68) |
| (0.1, 0.2), 2500 | 1190 | 94.9 | (2.19, 2.95) | 1190 | 95.0 | (2.12, 2.84) |

[a] $p_1$ and $p_2$ are marginal probabilities of capture in streams 1 and 2
